# TIDAL – Tool to Implement Developmental Analysis of Longitudinal data

**DOI:** 10.1101/2024.08.12.24311854

**Authors:** Alex S. F. Kwong, Amelia Edmondson-Stait, Eileen Xu, Ellen J. Thompson, Richard M. A. Parker, Ahmed Elhakeem, Liana Romaniuk, Rebecca M. Pearson, Kate Tilling, Thalia C. Eley, Andrew M McIntosh, Heather C. Whalley

**Author notes:** Denotes joint authorship (Alex S. F. Kwong, Amelia Edmondson-Stait).

## Abstract

**Motivation:** Growth curve modelling is one method used to model trajectories of traits and behaviours over time. However, accessing, analysing and interpreting trajectories requires statistical expertise, thereby creating potential barriers for users to implement and understand longitudinal traits. TIDAL is a user-friendly research tool designed to facilitate trajectory modelling by improving access, analysis and interpretation of trajectory and longitudinal data.

**Implementation:** TIDAL is available in two formats: an R package and an online Shiny application. The R package can be used offline, negating the need to upload potentially sensitive data.

**General features:** TIDAL includes all the main steps of trajectory analysis including: 1) data preparation, (converting data from wide to long format); 2) data exploration, via basic plots and descriptive information; 3) analysis of trajectories using mixed effects modelling, interpretation of results, visualisation of trajectories, and extraction of key features (scores at different ages; area under the curve); and 4) interactions to derive population specific trajectories, combined with all the above. TIDAL is built with a simple graphical interface to guide users through each step. R syntax accompanies each step.

**Availability:** Both versions of TIDAL can be found here: [https://tidal-modelling.github.io/].

## Introduction

Growth-curve modelling uses repeatedly-measured data from the same individuals to estimate trajectories that describe how variables change over time, identify key periods of change and why they occur.^1,2^ This method is used in life-course epidemiology and related disciplines including psychology, social science and public health.^3,4^ Of particular interest is examining trajectories according to key potential effect modifiers. Some examples include estimating trajectories of height and how they vary by sex,^5^ body mass index (BMI) trajectories stratified by levels of social adversity,^6^ cross cohort differences in bone mineral content (BMC) trajectories^7^ and depression trajectories stratified by genetic risk.^8^

Multi-level growth curve modelling (MLM; also known as hierarchical modelling or mixed-effects modelling)^9,10^ is one method for estimating trajectories. It takes its name from the fact that longitudinal data have a hierarchical or multilevel structure, with repeated observations (i.e., of height) nested within individuals.^2^ This nesting means the observations are not independent and analyses which ignore this clustering can result in inappropriate standard errors and potentially erroneous results. MLM is a popular and flexible method that can be applied to individuals throughout the life course.^4^ Leveraging a recent influx of longitudinal data and applying growth curve methods could enhance our understanding of longitudinal traits and behaviours across development potentially leading to better interventions and preventions.

However, there are significant barriers in place that prohibit the successful use and implementation of trajectory modelling. Whilst most statistical software packages accommodate growth curve analysis including Stata, R, SPSS, Python and SAS, they are not all accessible to users and many require a degree of time, costs and expertise to successfully implement and interpret. For example, users without statistical backgrounds may find it difficult to use some statistical software or not have capacity to learn new methods (time or costs to train), despite in depth knowledge of the trait they are interested in. Additionally, whilst some applications and resources exist that facilitate trajectory modelling,^11,12^ they all require statistical and software expertise to successfully implement and are not necessarily accessible and interpretable for naïve users. Finally, and perhaps more importantly, even experienced users may struggle to interpret more complex trajectories, which is especially the case with non-linear trajectories that often result in estimates that are not easily interpretable.^13^

To address these barriers, we have developed TIDAL (Tool to Implement Developmental Analysis of Longitudinal data), a free and easily accessible research tool for data wrangling, analysing, visualising and interpreting growth curve model data aimed at a variety of users, including researchers, clinicians, public health officials, government analysts and educators.

### Implementation

The TIDAL application can be found here: [https://tidal-modelling.github.io]. The tool is available in two formats: an R package and an online RShiny application. The R package and can be downloaded and installed locally and used offline, allowing users to analyse potentially sensitive data locally without uploading data to online servers.

The TIDAL website contains an overview of the tool, five example datasets, detailed instructions and a guided tutorial on how to install, run and interpret TIDAL (in video and PDF formats), a frequently asked questions section and some guidance for further information on statistics and growth curve modelling.

We have developed TIDAL to guide users through the main steps involved in growth curve modelling analysis. These include:

*1. Data import and preparation:* the first stage of TIDAL is to import the data. TIDAL accepts comma separated .csv or tab delimited .txt or .tsv files to the limit of 400MB. These data would typically be continuous variables such as BMI assessments across 4 occasions, with corresponding age and covariate information, or mental health scores across 5 waves split by treatment arms. Missing values should be set to “NA” or empty cells (“”). A synthetic dataset is embedded within the tool for exploratory use and analysis if required. TIDAL will convert data from wide (one row for each individual) to long (one row for each measurement occasion for each individual) format to accommodate multilevel growth curve modelling. This step can be bypassed if the user already has a long format dataset.

If the user imports wide format data, then they are first required to select identifier/subject, repeated ages/times/occasions and repeated outcomes variables from the dropdown boxes provided (see Figure 1 for an example). Variables are then created for a new long format dataset to be analysed, and users can rename these from the default to more relevant names. A snapshot of this new long format data is presented for users to ensure their data has been formatted correctly with error messages appearing if an unequal number of occasions and outcome variables have been entered (i.e., four occasion rows but only three outcome rows). Users can download the newly formatted long dataset for analysis outside of TIDAL if desired.

**Figure 1.**
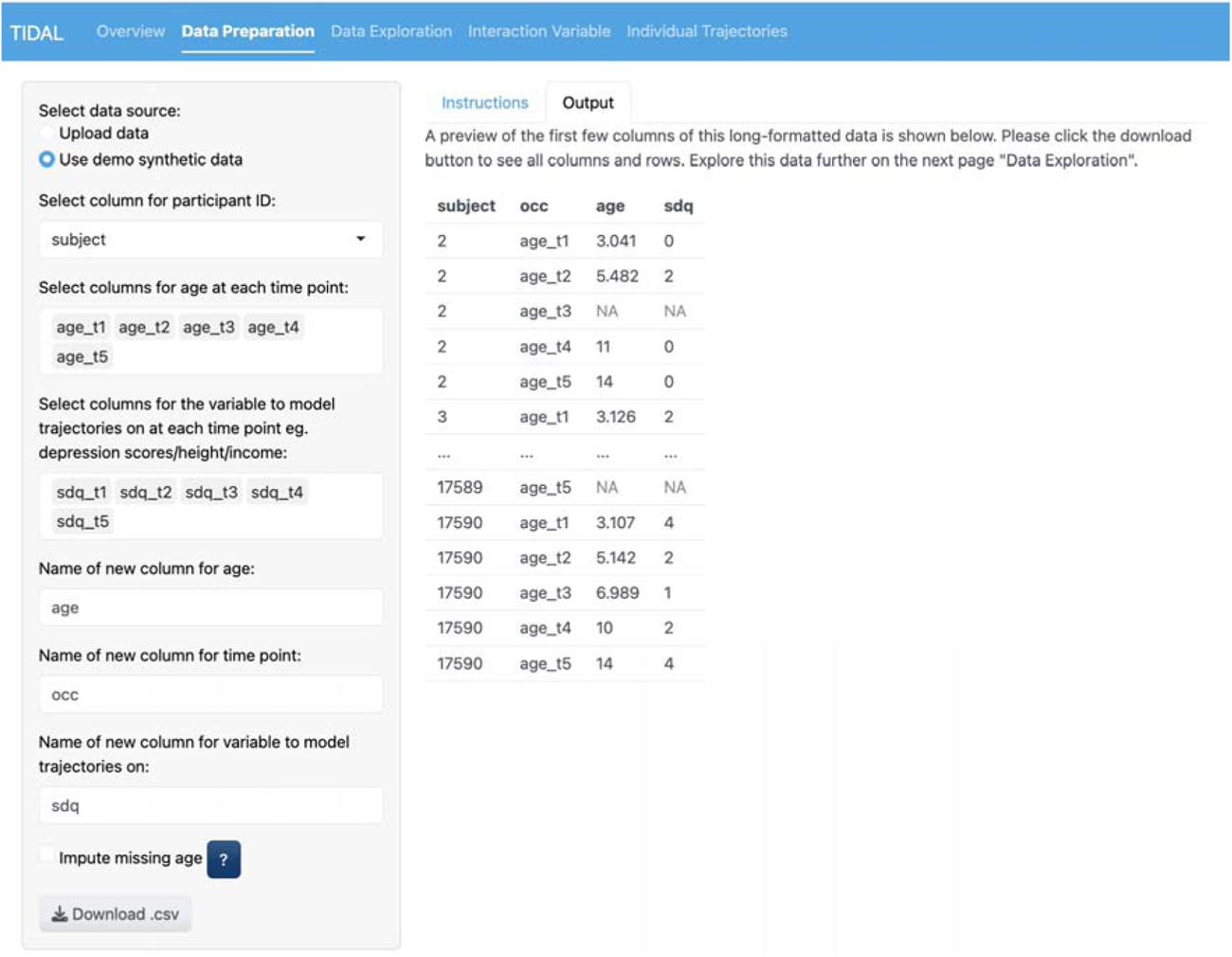
The data preparation stage for TIDAL. This figure shows the data preparation stage using the built in example data in TIDAL. The first three dropdown options include the variables subject, age_t1 to age_t5 and sdq_t1 to sdq_t5, which are selected from the original wide dataset. The bottom three dropdown options can all be renamed. Here the defaults of time_point and score have been renamed to occ and sdq, respectively. On the right, a table under the options tab shows the new data structure in long format.

*2. Data exploration and analysis:* the next stage of TIDAL uses the newly formatted long dataset from the previous stage or a new user uploaded long dataset to begin exploring the data. Users are required to select the relevant names from the dropdown boxes corresponding to the identifier/subject, outcome and age/time/occasion variables respectively (see Figure 2).

Users can then select the type of growth curve model from a choice of linear and non-linear functions to describe the association of the outcome with time/age and can specify fixed (population) and random (individual) effects. The lme4 package in R is used to conduct the growth curve modelling.^14^ Further information regarding the statistical properties of these models is given in the supplement. Currently, TIDAL supports trajectory modelling using fixed linear, quadratic, cubic and quartic polynomial terms, and random intercept, linear and quadratic terms (thereby allowing random slopes), as per previous research.^1,7,15^ To guide the choice of modelling, a line plot of the mean scores at each occasion is presented, alongside descriptive information (N, mean, standard deviation, median and interquartile range of the outcome variable at each occasion). Users need to select a model to initiate the descriptive information and plot, so we recommend running a linear model to begin exploring the data. Users can choose to include continuous or categorical covariates and survey weights into their models.

The time taken to run a model will depend on several factors. Models with many participants (more than 10,000), many repeated assessments (more than 5), complex missing data patterns or an increasing number of fixed and/or random effects will result in greater model complexity and longer waiting times. However, most models should complete in under 30 seconds based upon our testing across datasets and traits. Should the model not complete or fail to converge, error messages highlighting the underlying issue will appear.

Once the model completes, the lme4 syntax used to run the model in R will be presented. Users could adapt this syntax outside of TIDAL to create more complex models. The model estimates are presented giving the effect sizes, 95% confidence intervals and P values, alongside a value for model fit (deviance). A table of random effect estimates is also presented. Importantly, interpretation of all these results is given to aid understanding. This interpretation has been co-produced with users and lived experience to ensure the best possible interpretation of results for broad audience. Users are then directed to the “Plot” tab to visualise the trajectories with 95% confidence intervals (plotted alongside the mean scores from each occasion if desired). To further aid interpretation, we have also included the ability to calculate the outcome at a given age/time along the trajectory (i.e., depression scores at ages 13, 15 or 17), which is especially helpful for interpreting non-linear models with complex and potentially contrasting polynomial terms.^15^ Finally, we also include a feature which calculates the area under the curve (AUC), which represents the accumulation or cumulative exposure the population spends with the given trait or behaviour (i.e., how long on average individuals spend with depression or BMI). Users can then download a summary report that includes all this output. Statistical information supporting these methods are given in the supplement and on the GitHub.

**Figure 2.**
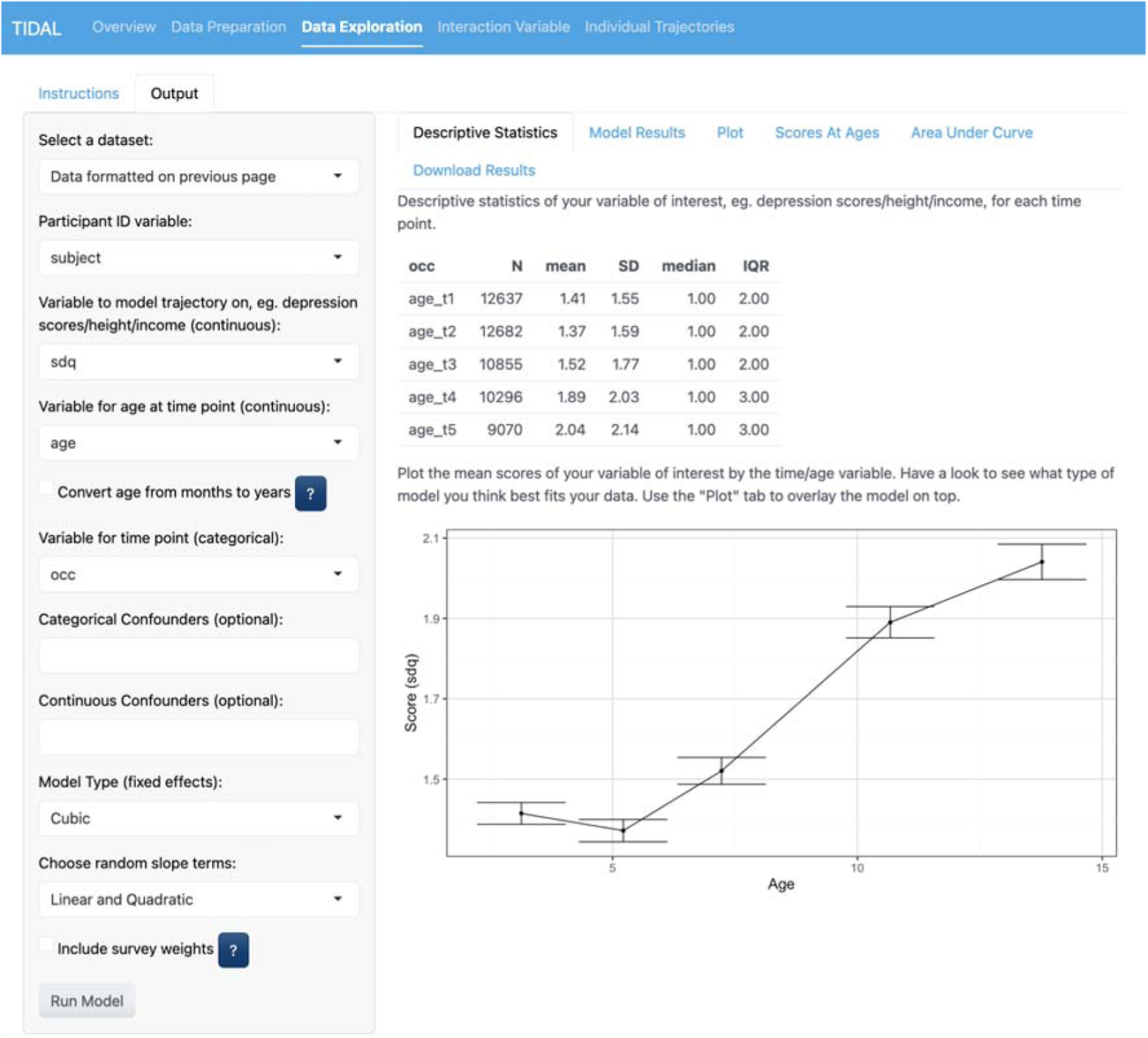
The data exploration and analysis stage of TIDAL. This figure shows the data exploration stage using the formatted data from the previous stage (highlighted by the first drop box). The variables sdq, age and occ are all selected as outcome and time variables respectively, carried forward from the previous stage. The right side shows the descriptive statistics and a plot of mean scores by age/time to guide the choice of trajectory modelling. In this case, the data appears to represent a non-linear trajectory, and a cubic model is chosen from the Model Type dropdown box with random linear and quadratic slope terms.

*3. Interactions:* users can obtain group/population specific trajectories by using the interactions tab and select between continuous and categorical variables on which to base the interactions. Continuous variables are standardised to have a mean of zero and a standard deviation of 1, whereas categorical variables will use the level with the lowest score as the reference group. The user is then presented with all the features stated above, but specific to interactions. Additionally, the user can make statistical comparisons between groups at given ages/times (i.e., height differences at age 12 between females and males) and between groups for the AUC (i.e., whether people with lower SES spend longer with greater depressive symptoms).

*4. Individual level trajectories:* finally, users can explore individual participant trajectories and observe how they vary compared to the overall population or groups/populations from the model. Users can plot up to 30 random individual trajectories from the overall population or specific to each population (i.e., 30 male trajectories and 30 female trajectories) or a specific subset of trajectories (taken from the corresponding identifier/subject variable).

### Use

We show how TIDAL can be used within the R package, although the implementation above is the same across both formats. TIDAL can be installed using the following commands in R.

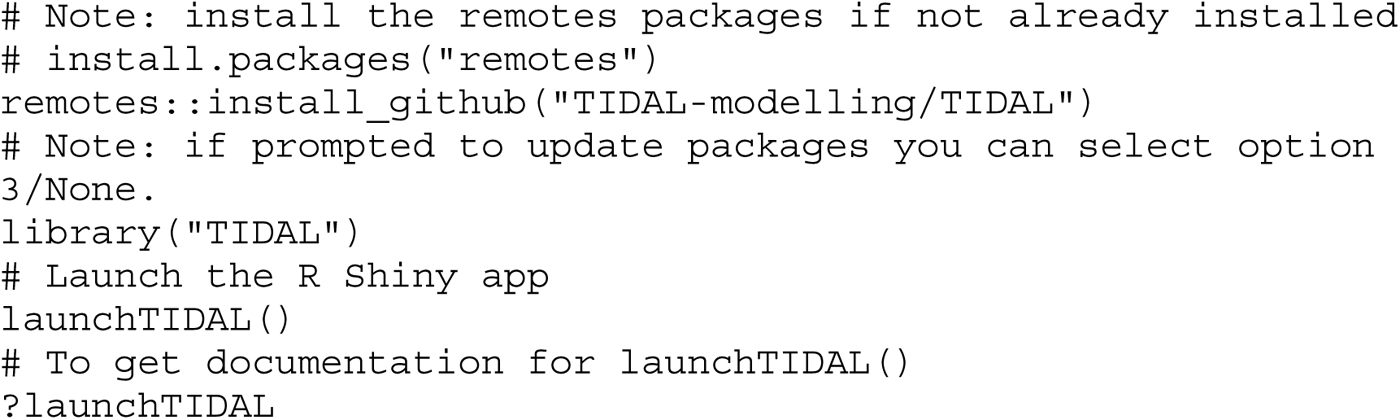

We highlight two examples of how TIDAL can be implemented using synthetic data, which holds the same properties as real data, but without concerns over privacy.^16^ The datasets used in this example can be found here: [https://tidal-modelling.github.io/docs/installation/synthetic_data.html]. Further information about both cohorts is given in the supplement.

#### 1. Trajectories of emotional symptoms across childhood and adolescence

The dataset in this first example comes from the Millenium Cohort Study^17^, and contains 12,720 individuals with up to 5 repeated assessments of parent reported emotional symptoms (referred to as sdq from here) between the ages of around 3 (t1) and 14 (t5).^18^ The research question here is: how might emotional symptoms develop across childhood and through adolescence?

As shown in Figure 1, the variables subject (the individual’s study ID), age_t1-age_t5 (ages at each occasion between occasion 1 and 5) and sdq_t1-sdq_t5 (sdq scores at each occasion between occasion 1 and 5) are in the original wide dataset and made into long format for analysis in TIDAL. Figure 2 then shows the variables subject, age, occ (occasion) and sdq are carried forward from the previous stage. No confounders or covariates are included in this model. After exploring a linear model first, it appears a cubic model may suit the data best given the non-linear pattern, so a cubic polynomial model with random intercept, linear and quadratic terms is selected. Figure 3 shows the output from the analysis stage of TIDAL, including the lme4 syntax for the model and information about the number of observations and people. Next, the fixed and random effects are presented, showing modest effect sizes for all fixed age polynomial terms. Interpretation of these results is then given by TIDAL as following:

> *“The score at the intercept is 1.56. The intercept here has been shifted to the mean age of all the assessments which is 7.58. You could interpret this as the mean score at the mean age (7.58) is 1.56.*

> *Every unit increase in age is associated with an increase of sdq by 0.10. As you have specified a cubic model, every unit increase in age∧2 is also associated with an increase of sdq by 0.007. Furthermore, every unit increase in age∧3 is also associated with a decrease of sdq by 0.002. However, it can be difficult to interpret these three estimates in isolation, so we would recommend exploring your trajectories with the “Plot” and “Scores At Ages” tabs for more information*.

> *The model fit (deviance) is 207821.889, you can compare this value to other similar models to determine which model has a better fit.”*

**Figure 3.**
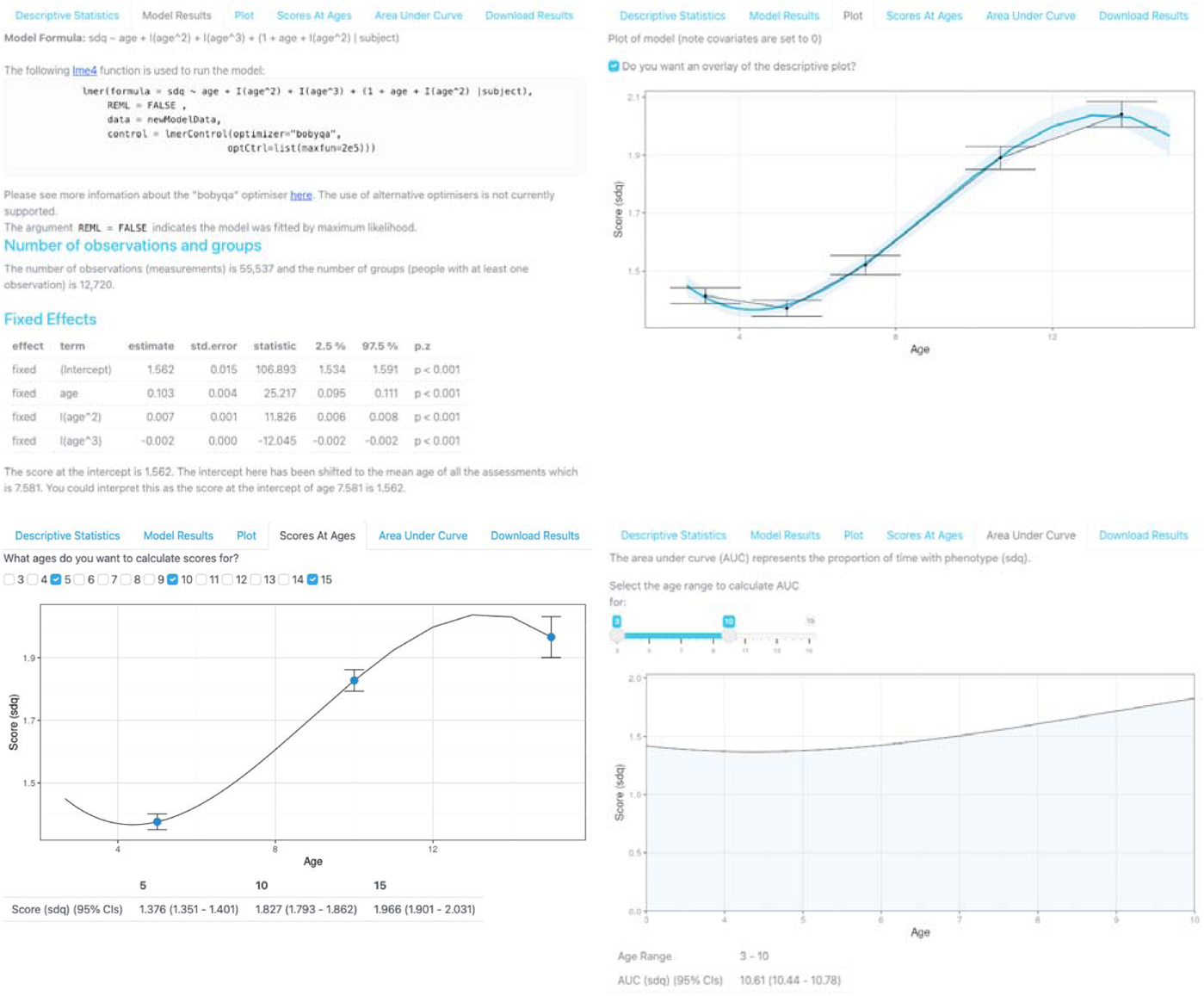
The data exploration and analysis stage of TIDAL continued. This figure shows the output from the corresponding model from Figure 2. Top left: the lme4 syntax used to run the model, followed by information about the number of observations and people included in the analysis and then the table of fixed effects and interpretation of the results. Top right: the plotted sdq trajectory with 95% CI and the descriptive plot from Figure 2 beneath. Bottom left: the calculation of scores at different ages, with the score for the sdq calculated at ages 5, 10 and 15 shown, alongside 95% CI. Bottom right: the area under the curve (AUC) plotted and calculated for sdq trajectories between the ages of 3 and 10.

Figure 3 also shows the predicted trajectories, overlaid onto the descriptive statistics plot. The model appears to match the observed descriptive data. To further aid interpretation, TIDAL also calculates sdq scores across all ages at which age data are available (i.e., between 3 and 15 here). We can see that the predicted score at age 5 is 1.37 (95%CI: 1.35, 1.40), at age 10 is 1.82 (95%CI: 1.79, 1.86) and at age 15 is 1.96 (95%CI: 1.90, 2.03). Finally, TIDAL has calculated the AUC, which in this example is restricted to ages 3-10 and shows that this population has an AUC score of 10.61 (95%CI: 10.44, 10.78). Together, these analyses suggest emotional symptoms scores tend to get higher with age, supporting similar research.^15^

#### 2. Sex differences in height from childhood to young adulthood

This next example uses data from the Avon Longitudinal Study of Parents and Children,^19^ and contains 10,261 individuals with up to 8 repeated assessments of height between the ages of around 7 and 20. The research question here is how do height trajectories vary between girls and boys across childhood and adolescence?

For brevity, we only demonstrate analysis using the interactions tab shown in Figure 4, as the process up to that point will be identical to the above. Figure 4 shows large effect sizes for the fixed age terms, and modest effects for the main effect of sex (variable name is female) and the interactions between sex and age terms. Interpretation of these results is then given by TIDAL:

> *“The interaction variable you have chosen has been factorised with the lowest level “female0” being the reference or baseline category. For the level “female0”, the score at the intercept is 153.04. The intercept here has been shifted to the mean age of all the assessments which is 12.02. You could interpret this as the mean score for people with “female0” at the average age (12.02) is 153.04*.

> *For the level “female0”, every unit increase in age is associated with an increase of “height” by 5.65*.

> *As you have specified a quadratic model, every unit increase in age∧2 is also associated with a decrease of height by 0.11. To estimate the effect of different trajectory groups, you can add the intercept, age and age∧ estimates to the corresponding interactions, age:interactions and age∧2:interactions to get group specific trajectories*.

> *However, it can be difficult to interpret these estimates in isolation, so we would recommend exploring your trajectories with the “Plot” and “Scores At Ages” tabs for more information.”*

**Figure 4.**
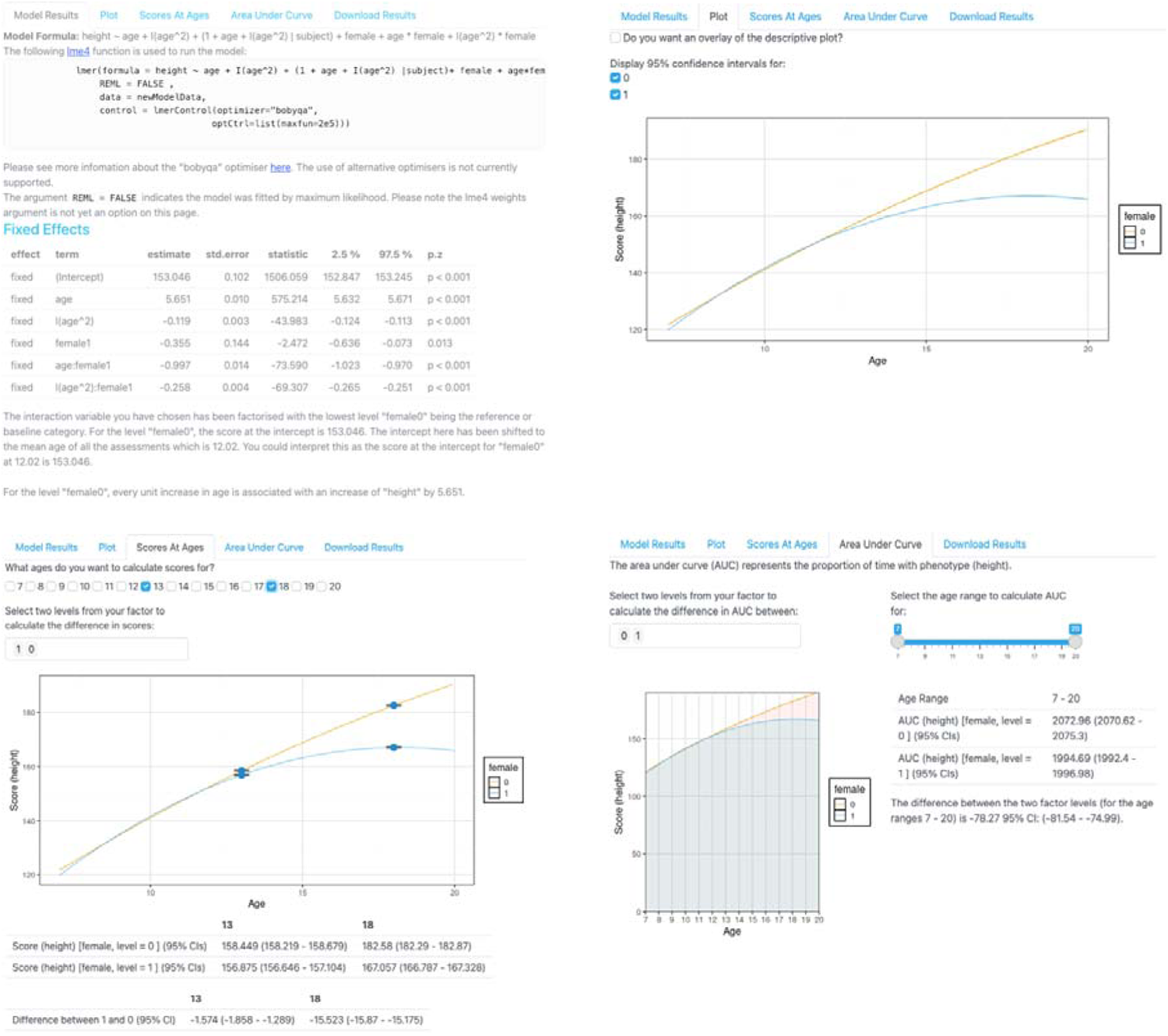
The interaction stage of TIDAL. This figure shows the output from the height trajetcories model. Top left: the lme4 syntax used to run the model, followed by information about the number of observations and people included in the analysis and then the table of fixed effects and interpretation of the interaction results. Top right: the plotted height trajectories of girls (1) and boys (0) with 95% CI. Bottom left: the calculation of height at different ages, with the differences in height calculated for ages 13 and 18 shown, alongside 95% CI. Bottom right: the area under the curve (AUC) plotted and calculated for height trajectories between the girls and boys between the ages of 7 and 20, accompanied by the AUC score and 95% CI.

Figure 4 shows the predicted trajectories, split by sex. Again, to further aid interpretation, TIDAL has also calculated height across all the ages at which age data is available (i.e., between 7 and 20). We can see that the predicted height at age 13 for boys is 158.44cm (95%CI: 158.21, 158.67) and at age 18 for boys is 182.58cm (95%CI: 182.29, 182.87). Whilst for girls, height at those ages is 156.87cm (95%CI: 156.64, 157.10) and 167.05cm (95%CI: 167.78, 167.32), respectively. TIDAL will also calculate the difference between girls and boys at those age ranges, giving a difference of -1.57cm (95%CI: -1.85, -1.28) at age 13 and a difference of -15.52cm (95%CI: -15.87, -15.17) at age 18. Finally, TIDAL will calculate the AUC for both girls and boys, with the difference in height AUC being - 78.27 (95%CI: -81.54, -74.99). Together, results show that girls and boys have similar height trajectories until about the age of 12 or 13, when boys continue to grow into young adulthood and girls tend to plateau in height, as demonstrated in previous research.^20^

## Discussion

TIDAL (Tool to Implement Developmental Analysis of Longitudinal data) is a research tool that facilitates access, analysis and interpretation of trajectory data. TIDAL comes in two formats, allowing users to go from raw data to visualised and interpretable trajectories in a few simple steps. A main aim of TIDAL is to remove some of the existing barriers in examining and understanding longitudinal traits and is aimed particularly at researchers, clinicians, public health experts, government analysts and educators. Our hope is that TIDAL allows users without experience in longitudinal modelling or those without capacity to train, to make progress in understanding longitudinal traits that could develop better interventions and preventions for health conditions and wellbeing.

Longitudinal studies are becoming more essential within research. This brings about challenges with how to appropriately analyse this data, especially for more complex longitudinal patterns. One of the main benefits of TIDAL is the interpretation of complex trajectories, particularly non-linear trajectories. The ability to visualise and extract information from trajectories (i.e., scores at different ages/times, AUC) is pertinent for understanding more about traits or behaviours of interest. An additional benefit of TIDAL is the ability to build upon the provided lme4 syntax for analysis outside of TIDAL for more complex models. It is worth noting that TIDAL only uses polynomial terms to handle time/age and additional functions such as splines,^5^ and fractional polynomials,^6^ may better suit the data. Future versions of TIDAL will look to include these functions of time, alongside additional features such as the age of peak velocity or age of peak symptoms.^1^ Two further offline desktop versions (Windows and macOS) are also in development. Likewise, latent class growth analysis or growth mixture modelling may be a better to use method depending on the research question.^21^

Like all research tools, TIDAL should be used responsibly and cannot mask poorly designed or inappropriate studies. We have tested TIDAL in seven longitudinal studies (including UK Biobank and Twins Early Development Study) with different outcomes and developmental stages and found it to be robust across these datasets. However, issues may arise in studies with substantial study attrition, as in other software packages. Users should have some experience or understanding of the data they are using as this will help ensure TIDAL runs successfully and the results are robust. It should also be noted that TIDAL uses the lme4 package and so is limited by the power of the original R package. Alternative software is available (i.e., MLwiN) to fit multilevel models, which vary in estimation method and the model specifications they can accommodate, with the caveat that these require statistical experience and costs if outside the UK.

In conclusion, TIDAL is a user-friendly research tool designed to help users access, analyse and interpret trajectories data. TIDAL has all the features that users would find in published manuscripts and goes beyond much of the existing research to extract pertinent information from trajectories. We hope that TIDAL will benefit prospective users of longitudinal data and aid interpretation and understanding of longitudinal traits.

## Data Availability

Please note that the study website contains details of all the data that is available through a fully searchable data dictionary and variable search tool" at the following website: http://www.bristol.ac.uk/alspac/researchers/our-data/.
Further information about MCS can be found here: https://cls.ucl.ac.uk/cls-studies/millennium-cohort-study/.

## Funding

The UK Medical Research Council and Wellcome (Grant ref: 217065/Z/19/Z) and the University of Bristol provide core support for ALSPAC. A comprehensive list of grants funding is available on the ALSPAC website: http://www.bristol.ac.uk/alspac/external/documents/grant-acknowledgements.pdf.

This publication is the work of the authors and ASFK will serve as guarantor for the contents of this paper. This research was funded by the Wellcome Trust through a Wellcome Mental Health Data Prize (Grant ref: 226686/Z/22/Z). For the purpose of Open Access, the author has applied a CC BY public copyright licence to any Author Accepted Manuscript version arising from this submission. ASFK is supported by a Wellcome Early Career Award (Grant ref: 227063/Z/23/Z). AES is funded by the Wellcome Trust (Grant No. 108890/Z/15/Z).

## Acknowledgments

We are grateful for the entirely voluntary co-operation of the children who form the Millennium Birth Cohort and their mothers, fathers and other family members, and to the Centre for Longitudinal Studies (CLS), UCL Social Research Institute, for the use of the Millennium Cohort Study data and to the UK Data Service for making them available. However, neither CLS nor the UK Data Service bear any responsibility for the analysis or interpretation of these data.

We are also extremely grateful to all the ALSPAC families who took part in this study, the midwives for their help in recruiting them, and the whole ALSPAC team, which includes interviewers, computer and laboratory technicians, clerical workers, research scientists, volunteers, managers, receptionists and nurses. We would also like to acknowledge those who have given feedback to our tool, including clinicians, teachers, researchers and our young people advisory group and facilitators.

## Author Contributions

Creation, testing and development of the TIDAL app: AES, ASFK, EX, EJT. Data analysis: ASFK. Study concept and design: all authors. Manuscript writing: ASFK. Interpretation of the data and critical revision and editing of the manuscript: all authors. All authors read and approved the final manuscript.

## Conflict of Interest

None

## Notes

### Competing Interest Statement

The authors have declared no competing interest.

### Author Declarations

The data collection for the MCS is approved by the UK National Health Service Research Ethics Committee. Written consent was obtained from all parents in the MCS at each survey (for MCS1, South West MREC [MREC/01/6/19]; for MCS2 and MCS3, London MREC [MREC/03/2/022, 05/MRE02/46]; for MCS4, Yorkshire MREC [07/MRE03/32]; for MCS5, Yorkshire and The Humber-Leeds East [11/YH/0203]; for MCS6, London MREC [13/LO/1786]; for MCS7, North East York [REC ref 17/NE/0341]). Ethical approval for the study was obtained from the ALSPAC Ethics and Law Committee and the Local Research Ethics Committees.

