## Supplement for "TIDAL – Tool to Implement Developmental Analysis of Longitudinal data"

**TIDAL – Tool to Implement Developmental Analysis of Longitudinal data Supplementary Materials**

**Information on datasets**

Millenium Cohort Study^1,2^

The Millennium Cohort Study (MCS) is a longitudinal birth cohort study, following a

nationally representative sample of approximately 19,000 people born between Sept 1, 2000, and Jan 11, 2002, in either England, Scotland, Wales, or Northern Ireland. During recruitment, efforts were made to ensure adequate representation through oversampling disadvantaged and ethnic minority populations. The data collection for the MCS is approved by the UK National Health Service Research Ethics Committee. Written consent was obtained from all parents in the MCS at each survey (for MCS1, South West MREC [MREC/01/6/19]; for MCS2 and MCS3, London MREC [MREC/03/2/022, 05/MRE02/46]; for MCS4, Yorkshire MREC [07/MRE03/32]; for MCS5, Yorkshire and The Humber-Leeds East [11/YH/0203]; for MCS6, London MREC [13/LO/1786]; for MCS7, North East–York [REC ref 17/NE/0341]). Further information about MCS can be found here: <https://cls.ucl.ac.uk/cls-studies/millennium-cohort-study/>.

Avon Longitudinal Study of Parents and Children^3-5^

Pregnant women resident in Avon, UK with expected dates of delivery between 1st April 1991 and 31^st^ December 1992 were invited to take part in the study. 20,248 pregnancies have been identified as being eligible and the initial number of pregnancies enrolled was 14,541. Of the initial pregnancies, there was a total of 14,676 foetuses, resulting in 14,062 live births and 13,988 children who were alive at 1 year of age. When the oldest children were approximately 7 years of age, an attempt was made to bolster the initial

sample with eligible cases who had failed to join the study originally. As a result, when considering variables collected from the age of seven onwards (and potentially abstracted from obstetric notes) there are data available for more than the 14,541 pregnancies mentioned above: The number of new pregnancies not in the initial sample (known as Phase I enrolment) that are currently represented in the released data and reflecting enrolment status at the age of 24 is 906, resulting in an additional 913 children being enrolled (456, 262 and 195 recruited during Phases II, III and IV respectively). The phases of enrolment are described in more detail in the cohort profile paper and its update. The total sample size for analyses using any data collected after the age of seven is therefore 15,447 pregnancies, resulting in 15,658 foetuses. Of these 14,901 children were alive at 1 year of age. Ethical approval for the study was obtained from the ALSPAC Ethics and Law Committee and the Local Research Ethics Committees.

Please note that the study website contains details of all the data that is available through a fully searchable data dictionary and variable search tool" at the following website: <http://www.bristol.ac.uk/alspac/researchers/our-data/>.

Study data were collected and managed using REDCap electronic data capture tools hosted at the University of Bristol.^6^ REDCap (Research Electronic Data Capture) is a secure, web-based software platform designed to support data capture for research studies.

**Statistical methods**

**Estimating trajectories**

As described in the manuscript, TIDAL uses the lme4 package to estimate trajectories.^7^ This is a linear mixed-effects model package. A simple trajectory using a linear polynomial model with a random intercept and random slope in TIDAL can be described using the following equation:

$$y_{ij}=\beta_{0}+\beta_{1}t_{ij}$$

$$+u_{0j}+u_{1j}t_{ij}+e_{ij}$$

where $y_{ij}$ is the score (emotional symptoms/height) and $t_{ij}$ is the age or time (centred around the mean age/time of all the assessments) for individual $j$ at occasion $i$. $\beta_{0}$ and $\beta_{1}$ are the fixed intercept and linear polynomial terms respectively, and $u_{0j}$ and $u_{1j}$ are the random intercept and linear terms, respectively. Finally, $e_{ij}$ is the occasion-specific residual.

The random effects are assumed multivariate normal distributed with zero mean vector and constant covariance matrix.

$$\left( \begin{aligned} u_{0j} \\ u_{1j} \end{aligned} \right) \sim N \left\{ \left( \begin{aligned} 0 \\ 0 \end{aligned} \right), \left( \begin{matrix} \sigma_{u0}^{2} & \\ \sigma_{u01} & \sigma_{u1}^{2} \end{matrix} \right) \right\}$$

These terms can be unclear, so we’ll use an example of a linear model in TIDAL substituting the estimates from the model into the equations above. Here, we run a simple linear fixed effects and a random intercept and random slope model using 5 sdq assessments embedded within TIDAL the synthetic dataset, with no covariates.

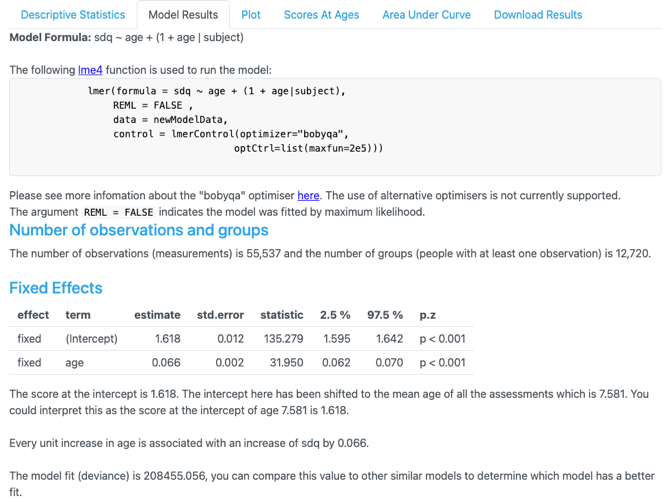

The fixed effects part of the linear model now looks like this to form the population trajectory:

$${sdq scores}_{ij}=1.618+0.066t_{ij}$$

Note that we do not include the random effects here ($u_{0j},u_{1j}$) as these are unique to each individual and would be used to create individual specific trajectories.

A more complex polynomial model using up to a quartic polynomial term with random intercept, linear and quadratic terms would take the following form:

$$y_{ij}=\beta_{0}+\beta_{1}t_{ij}+\beta_{2}t_{ij}^{2}+\beta_{3}t_{ij}^{3}+\beta_{4}t_{ij}^{4}$$

$$+u_{0j}+u_{1j}t_{ij}+u_{2j}t_{ij}^{2}+e_{ij}$$

where $y_{ij}$ is the score (emotional symptoms/height) and $t_{ij}$ is the age/time (centred around the mean age/time of all the assessments) for individual $j$ at occasion $i$. Then, $\beta_{0}$, $\beta_{1}$, $\beta_{2}$, $\beta_{3}$, and $\beta_{4}$ are the fixed intercept, linear, quadratic, cubic and quartic polynomial terms respectively, and $u_{0j}$, $u_{1j}$ and $u_{2j}$ are the random intercept, linear and quadratic terms, respectively. The occasion-specific residual variance is till specified by Finally, $e_{ij}$.

The random effects are again assumed multivariate normal distributed with zero mean vector and constant covariance matrix.

$$\left( \begin{aligned} u_{0j} \\ u_{1j} \\ u_{2j} \end{aligned} \right) \sim N \left\{ \left( \begin{aligned} \begin{aligned} 0 \\ 0 \end{aligned} \\ 0 \end{aligned} \right), \left( \begin{matrix} \sigma_{u0}^{2} & & \\ \sigma_{u01} & \sigma_{u1}^{2} & \\ \sigma_{u02} & \sigma_{u12} & \sigma_{u2}^{2} \end{matrix} \right) \right\}$$

Again, we can substitute estimates from TIDAL into the equation above using a quartic polynomial model as shown below.

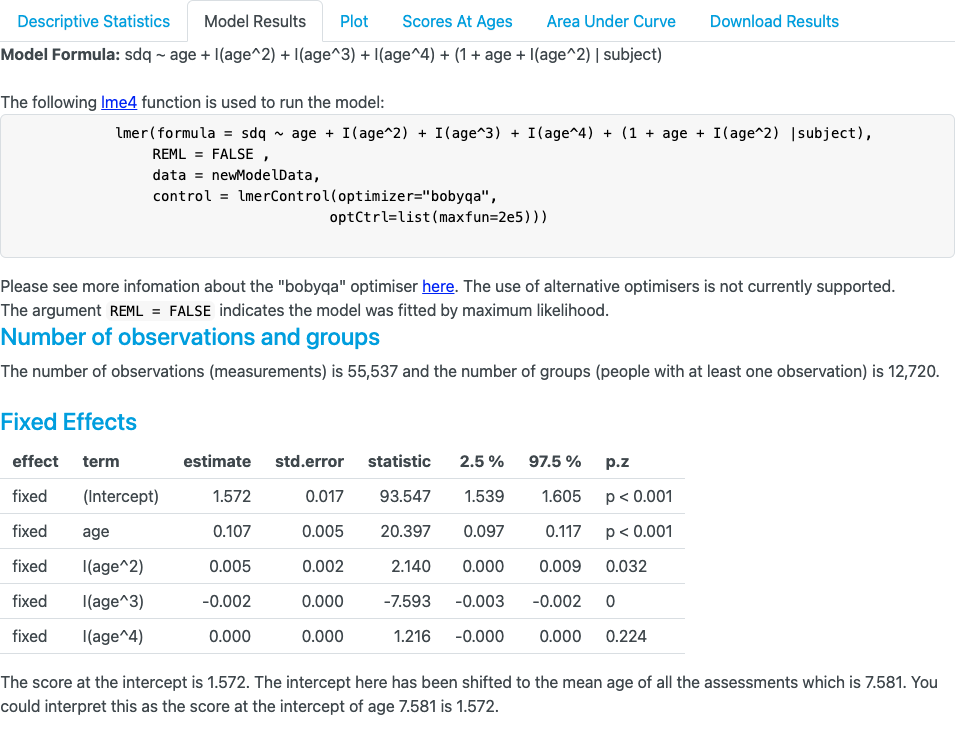

The fixed effects part of the quartic polynomial model now looks like this to form the population trajectory:

$${sdq scores}_{ij}=1.572+0.107t_{ij}+ 0.005t_{ij}^{2}+-0.002t_{ij}^{3}+0.0001t_{ij}^{4}$$

Again, note that we do not include the random effects here ($u_{0j},u_{1j},u_{2j},$) as these are unique to each individual and would be used to create individual specific trajectories.

An alternative way of parameterising these equations is shown below. For the linear model with a random intercept and random slope, as described above, this can be specified as the following:

$$E\left( y_{ij} | t_{ij} \right)= \beta_{0}+\beta_{1}t_{ij}$$

Individual specific trajectories would take the following form:

$$E\left( y_{ij} | t_{ij}, u_{0j}, u_{1j} \right)= \beta_{0}+\beta_{1}t_{ij}+ u_{0j}+ u_{1j}$$

**Interactions and population level trajectories**

We can now start to adapt the equations above to include interactions and population specific trajectories (i.e., trajectories of males and females). We can adapt the simple linear model described above to include a covariate for sex:

$$y_{ij}=\beta_{0}+\beta_{1}t_{ij}+\beta_{2}x_{1j}$$

$$+u_{0j}+u_{1j}t_{ij}+e_{ij}$$

where $\beta_{2}x_{1j}$ is the parameter that denotes being female ($x_{1j}$= 1) or male ($x_{1j}$= 0). Note this model would just adjust for sex in the model. To create an interaction or sex specific trajectory, we can interact this covariate for sex with the time variable ($\beta_{1}t_{ij}$) to create the following:

$$y_{ij}=\beta_{0}+\beta_{1}t_{ij}+\beta_{2}x_{1j}+\beta_{3}x_{1j}t_{ij}$$

$$+u_{0j}+u_{1j}t_{ij}+e_{ij}$$

where $\beta_{3}x_{1j}t_{ij}$ is the parameter for sex interacted with time. For males (where $x_{1j}=0$), $\beta_{2}x_{1j}\& \beta_{3}x_{1j}t_{ij}$ can be set to 0 as (as any parameter multiplied by 0 is 0), so the male trajectory would just be $y_{ij}=\beta_{0}+\beta_{1}t_{ij}$. For females, (where $x_{1j}=1$), you would include the parameters of $\beta_{2}x_{1j} \& \beta_{3}x_{1j}t_{ij}$, so the female trajectory would be the full equation of $y_{ij}=\beta_{0}+\beta_{1}t_{ij}+\beta_{2}x_{1j}+\beta_{3}x_{1j}t_{ij}$. Note, we do include random effects here. An alternative way of parameterising these population specific trajectories is as follows:

Male trajectory: $E\left( y_{ij} | t_{ij}, u_{0j}, u_{1j} \right)= \beta_{0}+\beta_{1}t_{ij}$
Female trajectory: $E\left( y_{ij} | t_{ij}, u_{0j}, u_{1j} \right)= \beta_{0}+\beta_{1}t_{ij}+\beta_{2}x_{1j}+\beta_{3}x_{1j}t_{ij}$

Using this framework and some estimates from TIDAL (again using 5 occasions of the SDQ data, with female [coded as 0 for males and 1 for females] interacted with the trajectories), the above equation can be shown as followed:

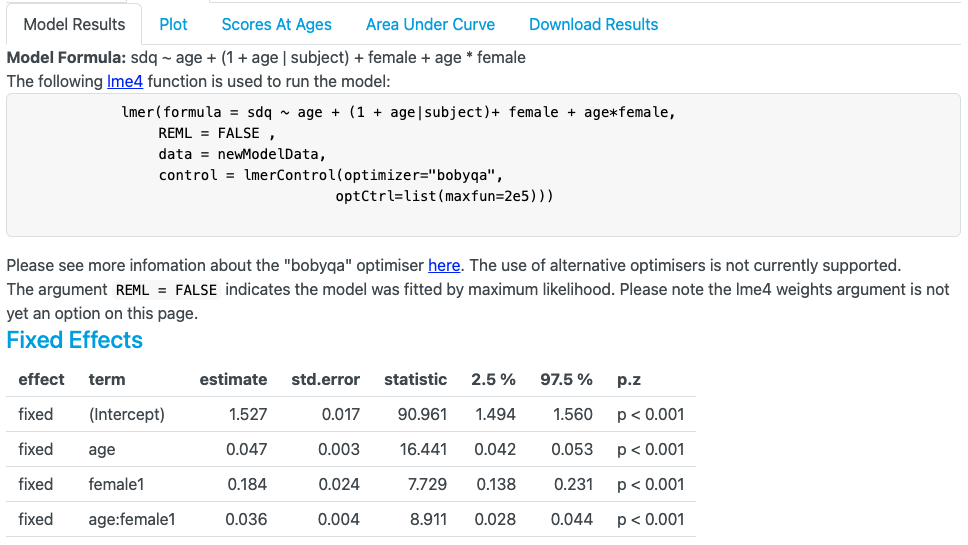

Male trajectory: $E\left( {sdq scores}_{ij} | t_{ij}, u_{0j}, u_{1j} \right)= 1.527+0.047t_{ij}$
Female trajectory: $E\left( {sdq scores}_{ij} | t_{ij}, u_{0j}, u_{1j} \right)= 1.527+0.047t_{ij}+0.184+0.036t_{ij}$

You could consider the male trajectory as being the baseline trajectory, with the effect of being female being added onto the baseline trajectory. If you have multiple levels within the covariate/interaction variable (i.e., if sex was coded as males = 0, females = 1, other = 2 or unknown = 3), then the interaction level 2 (other) estimates would be substituted in place of the female estimates. A greater discussion of this method is given in the supplement of this paper.^8^

**Calculating scores at ages**

When calculating the scores/trait at each time/age/wave, we can use the equations above and simply substitute in time/age/wave to calculate the score at that occasion.

For example, if we wanted to calculate the score at age for males, we could take the following equation:

Male trajectory: $E\left( {sdq}_{ij} | t_{ij}, u_{0j}, u_{1j} \right)= 1.527+0.047t_{ij}$

and substitute in age 10 for $t_{ij}$ (whilst noting age has been mean centered to the mean age of all the assessments [age 7.581 as shown in the figures above] so 10 – 7.581 = 2.419). We replace $t_{ij}$ with 2.419 so the equation becomes:

Male trajectory: $E\left( {sdq}_{age10j} \right)= 1.527+(0.047*2.419)$

Male trajectory: $E({sdq}_{age10j})= 1.64$

For females, we use the same equation as the section above:

Female trajectory: $E\left( {sdq}_{ij} | t_{ij}, u_{0j}, u_{1j} \right)= 1.527+0.047t_{ij}+0.184+0.036t_{ij}$

which when calculating the score at age 10, we can use the same rationale as mentioned above, but this time for females:

Female trajectory: $E\left( {sdq}_{age10j} \right)= 1.527+\left( 0.047*2.419 \right)+0.184+(0.036*2.419)$

Female trajectory: $E({sdq}_{age10j})= 1.91$

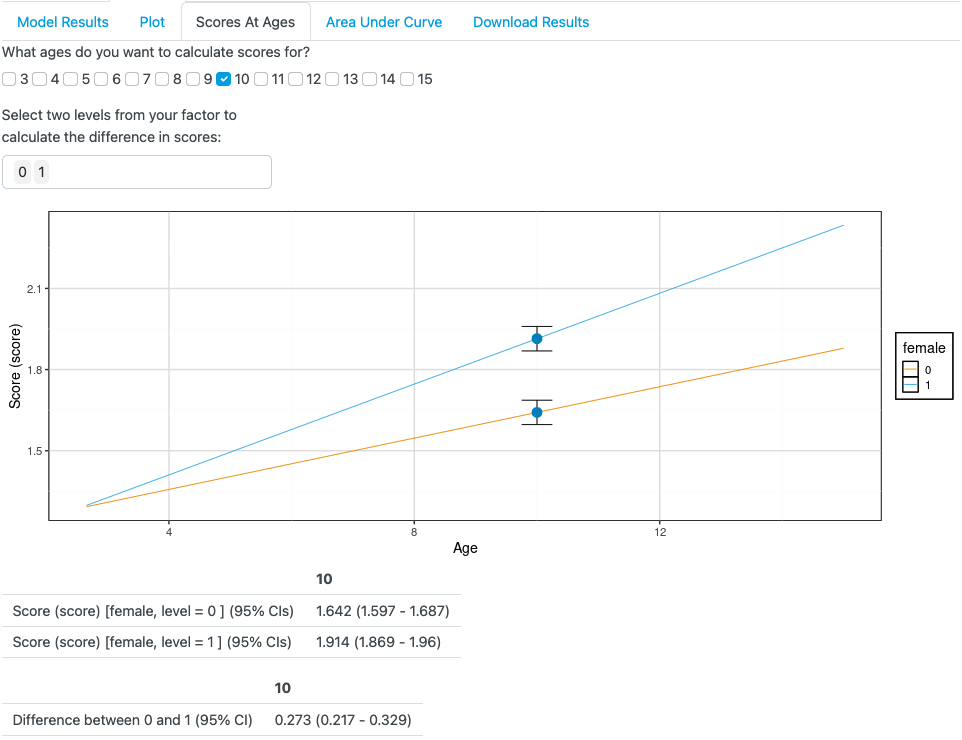
These scores match nicely to what is plotted in the figures. The ‘glht’ package is used to create confidence intervals and a linear comparison between the two groups. More information on how to calculate scores at more complex trajectories is given here.^8,9^

**Area under the curve**

To calculate the area under the curve (AUC), we first need to take the estimates from the model and pick ages/time to specify the period we want to examine (i.e., between the ages of 4 and 9 for example).

Here, we’ll use the estimates from a simple linear model in TIDAL using the 5 sdq occasions again. These results are shown below.

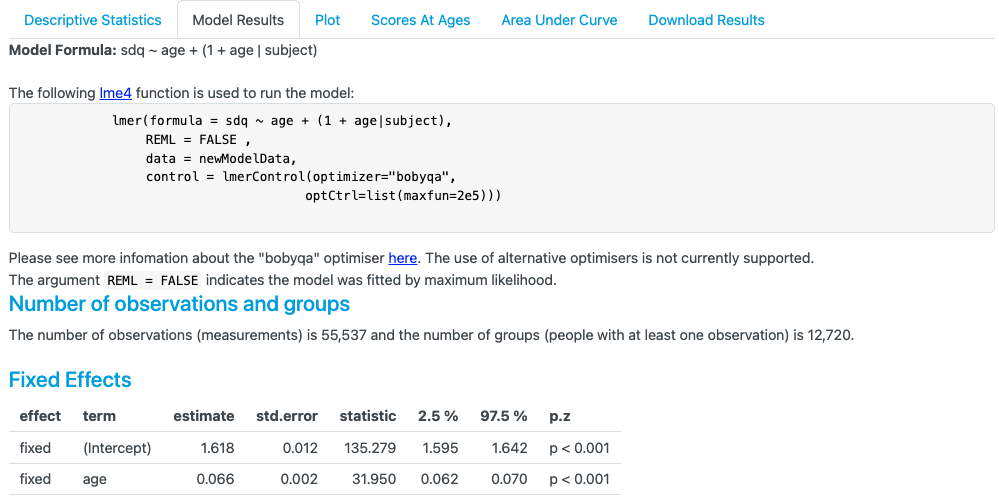

To calculate the AUC, we take the estimates from the model and first calculate the upper and lower bounds of the AUC period we want to estimate. The equation for a linear model will appear as follows:

Upper AUC score: $= \beta_{0}upperAUC+\frac{\beta_{1}\left( upperAUC \right)^{2}}{2}$

Lower AUC score: $= \beta_{0}lowerAUC+\frac{\beta_{1}\left( lowerAUC \right)^{2}}{2}$

To calculate the AUC between the ages 4 and 9. We can substitute these estimates and AUC values we want to calculate into the above equation (whilst noting age has been mean centered to the mean age of all the assessments [age 7.581 as shown in the figures above] so 9 – 7.581 = 1.419 and 4 – 7.581 is -3.581):

Age 9 AUC score: $= 1.618*1.419+\frac{0.066{*\left( 1.419 \right)}^{2}}{2}$

Age 4 AUC score: $= 1.618* -3.581+\frac{0.066{*\left( -3.581 \right)}^{2}}{2}$

Age 9 AUC score: $= 2.362$

Age 4 AUC score: $= -5.371$

The difference between these two scores (2.362 – (-5.371)) = 7.733, which we can see is calculated in the figure below. The ‘glht’ package is then used to create confidence intervals when calculating these estimates.

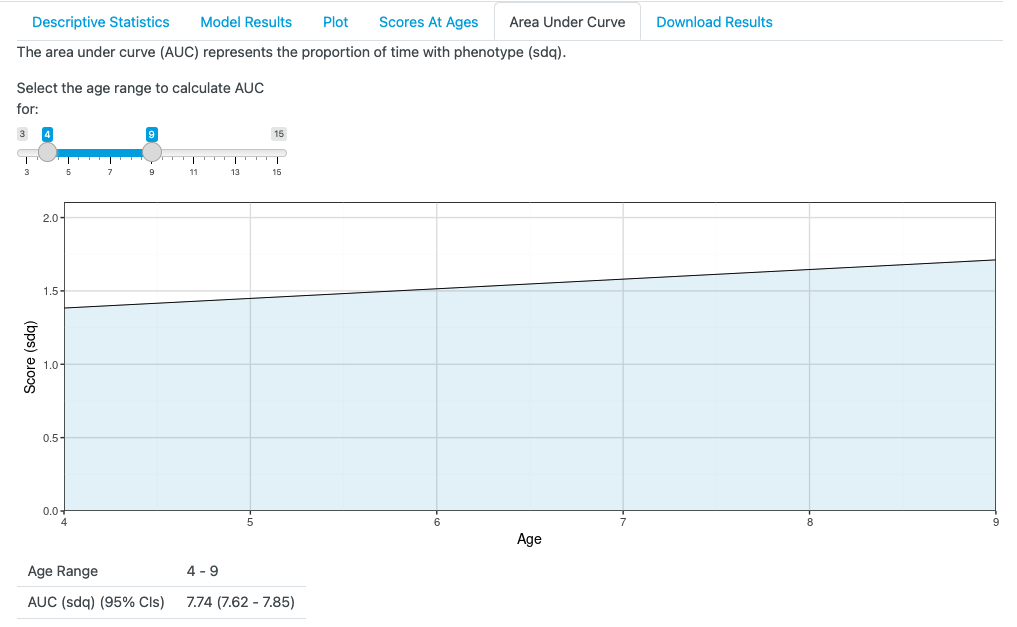

When calculating the AUC interactions, the same format as above can be used. For example, using a linear model and an interaction for sex (female coded as males =0, females = 1), we get the following estimates:

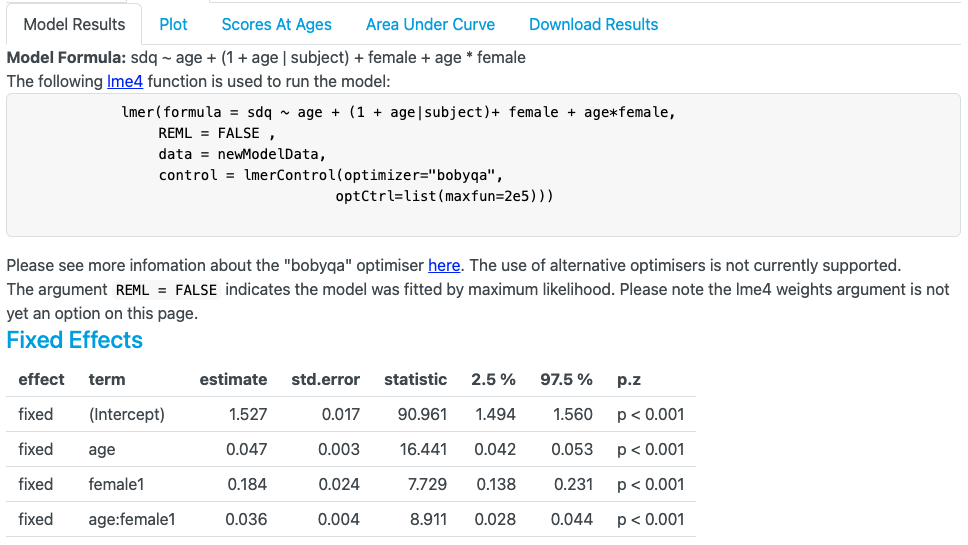

To calculate the AUC for males and females between the ages of 8 and 15, we could use the equations above (remembering that female estimates (female coded as 1) are added to the male estimates, and that age has been mean centered to the mean age of all the assessments [age 7.581 as shown in the figures above] so 15 – 7.581 = 7.419, and 8 – 7.581 is 0.419:

Male age 15 AUC: $= 1.527*7.419+\frac{0.047{*\left( 7.419 \right)}^{2}}{2}$

Male age 8 AUC: $= 1.527* 0.419+\frac{0.047{*\left( 0.419 \right)}^{2}}{2}$

Female age 15 AUC: $= 1.618*7.419+\frac{0.066{*\left( 7.419 \right)}^{2}}{2}+0.184*7.419+\frac{0.036{*\left( 7.419 \right)}^{2}}{2}$

Female age 8 AUC: $= 1.618*0.419+\frac{0.066{*\left( -3.581 \right)}^{2}}{2}+0.184*0.419+\frac{0.036{*\left( 0.419 \right)}^{2}}{2}$

Male age 15 AUC: $= 12.622$

Male age 8 AUC: $= 0.644$

Total male AUC between 15 and 8:$12.622-0.644=11.98$

Female age 15 AUC: $= 1.618*7.419+\frac{0.066{*\left( 7.419 \right)}^{2}}{2}+0.184*7.419+\frac{0.036{*\left( 7.419 \right)}^{2}}{2}$

Female age 8 AUC: $= 1.618*0.419+\frac{0.066{*\left( -3.581 \right)}^{2}}{2}+0.184*0.419+\frac{0.036{*\left( 0.419 \right)}^{2}}{2}$

Male age 15 AUC: $= 14.978$

Male age 8 AUC: $= 0.724$

Total female AUC between 15 and 8:$14.978-0.724=14.25$

The difference between the males AUC (11.98) and the females AUC (14.24) = 2.27, which matches the figure below, with differences due to rounding errors. Again, the ‘glht’ package is used to create confidence intervals and a linear comparison between the two groups.

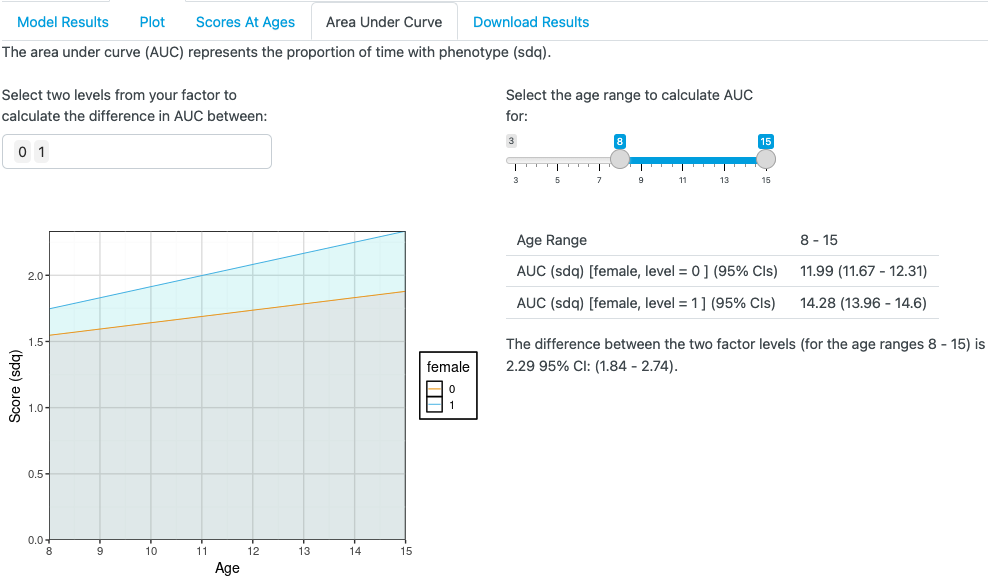
